# Modeling the flow of the COVID-19 in Germany: The efficacy of lockdowns and social behavior

**DOI:** 10.1101/2020.12.21.20248605

**Authors:** Muhamad Khairul Bahri

**Affiliations:** The Graduate Programme of Development Studies (Jurusan Pascasarjana Studi Pembangunan) Bandung Institute of Technology, 40132, Indonesia

**Keywords:** system dynamics, the COVID-19, the flow of the COVID-19, Germany COVID-19 cases, the SEIR model

## Abstract

This study develops a computer simulation in understanding the flow of the COVID-19 in Germany between January 2020 and July 2020. This aims to analyze not only the flow of the COVID-19 but also the efficacy of taken measures during the given period. The computer model is based on the SEIR concept and it is based on the system dynamics approach in which some uncertain parameters are estimated through the calibration process. Moreover, the SEIR computer model is developed by considering different flows of COVID-19 cases in older and young people in Germany. This study successfully reproduces similar patterns of infected, recovered, and death cases. Moreover, as the SEIR model can successfully reproduce similar patterns, the SEIR model can be a basis to estimate other resources such as health workers, and bed capacities.

## 1. Introduction

Originally, earlier cases of COVID-19 were reported from Wuhan, China in December 2019. Upon the international travel involving Chinese visitors, the COVID-19 has spread across the world, leading to the first pandemic in the last decade. Owing to its largest and widespread impacts, the WHO reports the latest figures on confirmed cases and deaths across the world since January 2020.

Germany, the largest economic producer in Europe, inevitably also has experienced this pandemic. The first COVID-19 case in Germany was reported in late January 2020 as the first confirmed patient contacted his infected colleague from China (e.g. Böhmer et al., 2020). Within two months, more than 100 confirmed cases were recorded in Germany (https://www.worldometers.info/coronavirus/country/germany/). Afterward, confirmed cases increased exponentially by about 6,000 in the mid-April 2020. By the end of July 2020, the COVID-19 cases decreased significantly as seen in figure 1.

**Figure 1.**
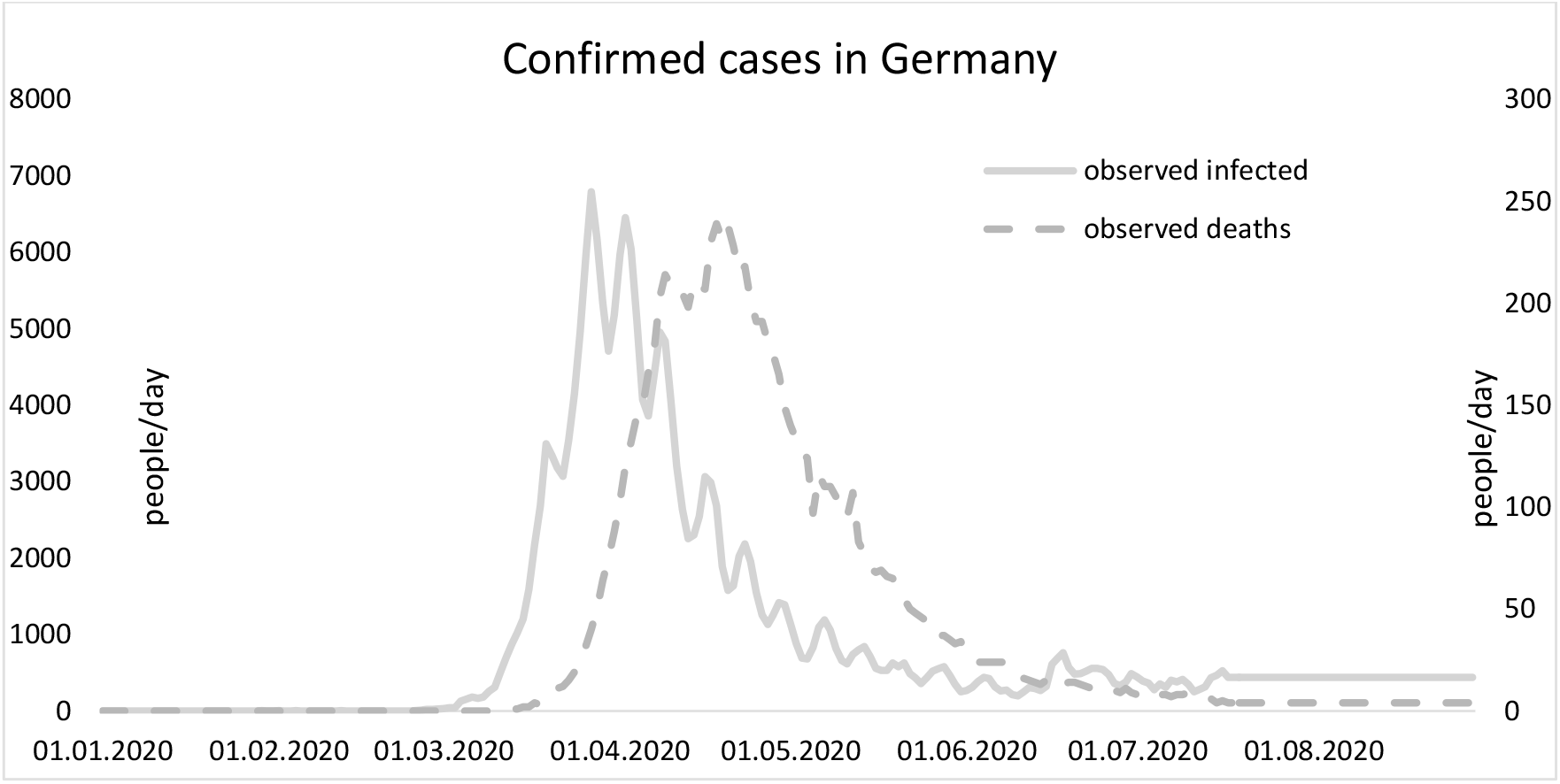
Confirmed infected and death cases in Germany

To anticipate the massive flow of the COVID-19, the federal government introduce public closures by closing public spaces such as schools, universities, and restaurants starting from March 16 (https://covid19-country-overviews.ecdc.europa.eu/#14_Germany). A further measure was also applied such as the national curfew ban and restricting people gathering. In principle, people were advised to stay at home as long as possible and leaving home only for basic needs.

Several studies analyzed the COVID-19 dynamics in Germany. Existing studies (Barbarossa et al., 2020; Khailaie et al., 2020) introduced mathematical modeling in understanding the dynamic flow of the COVID-19 while other existing studies (Dudel et al., 2020; Karagiannidis et al., 2020; Nachtigall et al., 2020) explained demographic data of patients. The flow of the COVID-19 also has been the main focus of existing studies (Böhmer et al., 2020; Stang et al., 2020).

Despite important contributions, no available study has explained any computer simulation to investigate the flow of the COVID-19 in Germany. Existing studies (Barbarossa et al., 2020; Khailaie et al., 2020) only explained the flow of the COVID-19 in Germany but they did not separate two different measures in tackling the flow of the COVID-19. This is important as existing studies (Lüdecke, & von dem Knesebeck, 2020; Hoenig, & Wenz, 2020) indicated two types of policies: behavior reduction policy (voluntarily personal actions) and lockdowns (the government policy with enforcement laws).

Hence, this study aims to develop a computer model that investigates the first wave of the COVID-19. Furthermore, no available study has analyzed two types of policies in slashing the pandemic flow and neither available study has developed the cohort SEIR model that is based on the SD approach. It is thus this study offers a computer model to explain the flow of the COVID-19 from January until July 2020 as well as analyzing the efficacy of the two different policies and consisting the cohort groups (young and old patients).

## 2. Methods and Data

The computer model is based on data collected by www.worldometer.com and Robert Koch Institute (RKI - www.rki.de). This study also data relate to important measures from https://covid19-country-overviews.ecdc.europa.eu/#14_Germany as this portal provides information step by step the federal government took measures in tackling the flow of the COVID-19. Data collection includes several data types such as infected, deaths, and recoveries. Other important data are also collected such as ages, incubation time, and recovery time.

The computer model follows the Susceptible, Exposed, Infected, and Recovered (SEIR) concept. This computer model is based on the system dynamics (SD) approach as a lot of studies have successfully simulated infectious, non-infectious diseases, and other healthcare issues (Donsimoni et al., 2020; Darabi & Hosseinichimeh, 2020; Davahli et al., 2020; Homer & Hirsch, 2006). While the computer model follows the SEIR concept and the SD approach, challenges owing to limited data such as uncertain parameters are visible. This study calibrates uncertain parameters using the calibration process available in Vensim©.

To do the calibration process, this study uses the Markov Chain Monte Carlo (MCMC) calibration process owing to unknown parameters. To obtain the best parameter values, the system dynamics model compares two outputs including infected cases and deaths. The two observed variables are compared with simulated variables to get the best parameter values in which means that the best parameter values lead to the smaller discrepancies between the two variables.

This study also introduces two measures or policies in tackling the flow of the COVID-19. The first policy is called behavioral risk reduction and the second one is lockdowns (public closures). The main reason to separate policies is the first policy is once infected people and disinfected people easy to identify, self-quarantines and health attitude such as hand washing are two possible solutions. However, once separating infected and disinfected people is relatively difficult owing to case spikes, the second policy i.e. lockdown is the best solution to stop the transmission of diseases (Piguillem & Shi, 2020). Separating these two policies is also important owing to reasons. The first reason existing studies explained the importance of the first policy (Lüdecke, & von dem Knesebeck, 2020; Hoenig, & Wenz, 2020) and the second reason is while the first policy is not supported by legal enforcement, the second policy is supported law enforcement (Hoenig, & Wenz, 2020).

## 3. Discussion and Results

The SEIR model consists of two groups: old (≥60 years) and young patients (<60 years). There is an imbalance proportion of the infected cases in the two groups as RKI (2020) has explained that the first group consists of about 95% of total deaths and about 21% of total infected cases. While the second group covers of 5% of total deaths and only 79% of total infected cases. This accommodates possible different factor patterns such as recovery time and infection duration between two groups.

Several studies (e.g. Karagiannidis et al., 2020; Nachtigall et al., 2020) pointed out that there is an imbalance effect of the COVID-19 in which older people tend to experience the most negative effects, especially high death cases among older people. It is thus separating patients in two groups means the cohort SEIR model represents imbalance effects of the COVID-19 across multiple age groups.

Table 1 displays the main variables used in this study and their estimated values based on existing studies. As previously mentioned, the MCMC calibration process is needed to obtain the best values, especially for unknown or uncertain parameters. During the MCMC calibration process, values of uncertain parameters are set based on table 1.

**Table 1.** The setting of parameter values for the calibration process

| Variables | Values | References |
| --- | --- | --- |
| The first confirmed case(s). | January 27<sup>th</sup> | (Böhmer et al., 2020; Stafford, 2020) |
| Ro (basic reproduction number) | (2.4-3.8) | (Khailaie et al., 2020; RKI, 2020) |
| Incubation time | (2-7) days | (Böhmer et al., 2020; Khailaie et al., 2020; RKI, 2020) |
| Infection duration | (45-91) days<br>This variable only occurs for older patients ($\geq 60$ years) as this group has deaths the most. | (Karagiannidis et al., 2020; Nachtigall et al., 2020; Stang et al., 2020) |
| Recovery time<br>* recovery time1<br>** recovery time2<br>Respectively, these variables represent recovery time for older patients ($\geq 60$ years) and younger patients | (3-35) days*<br>(3-7) days**<br>. | (Nachtigall et al., 2020; Karagiannidis et al., 2020) |
| Fraction based on age structure (fraction of group 1 compared with total infected patients) | (19% - 22%) | RKI (2020) |
| The earlier measures such as voluntary social distancing and handwashing behavior.<br>Two existing studies (Lüdecke, & von dem Knesebeck, 2020; Hoenig, & Wenz, 2020) explained detected protective behavior and public health campaign to minimize the COVID-19 flow around mid-February | Two complementing variables are “behavioral reaction time” (19-25 days) and “behavioral risk reduction” is assumed between 10% and 50%. The behavioral reduction time is measured between the first infection case and the beginning of the behavioral risk reduction actions. This time measurement also applies to other time measurements. | (ECDC, 2020; Der Spiegel, 2020) |
| The national lockdown (public closures).<br>This means that the national lockdown started (56-62) days prior to the first infection.<br>The national lockdown is a very strict measure so the effect is also assumed very high (60-85)% | Public closures starting from March 16<sup>th</sup> and banning more than 2 people on March 22<sup>th</sup>.<br>In the model, this variable is called “lockdown reaction time” (56-62), and its complement variable is called “lockdown risk reduction” (60-85)%.<br>As the lockdown starts may vary across the local authorities, another variable i.e. “delay time” (1-5 days) is inserted in the computer model. | (ECDC, 2020; Stafford, 2020) |

Figure 2 shows that the system dynamics model (SD) separates infected patients into two groups. This also means that for each group, the SD model calculates the number of infected cases, recoveries, and deaths. Recovery time1 is defined the average time between the symptom onset and recovery while infected duration1 is defined as the average time between the symptom onset and deaths.

**Figure 2.**
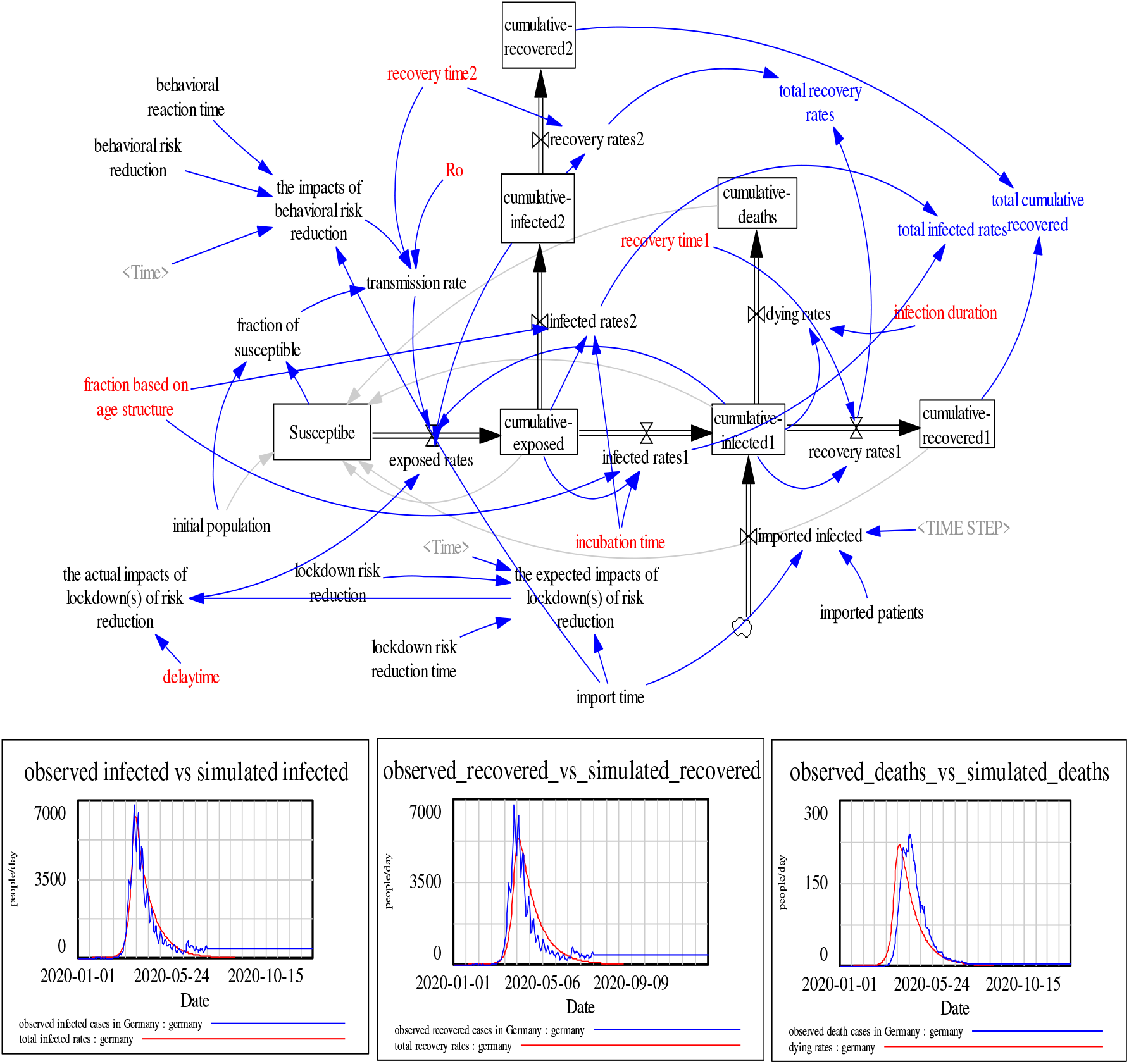
The SD model of Germany COVID-19

The number of infected cases, deaths, and recoveries are based on equations 1-3 (similar equations apply for *infected rates2* and *recovery rates2*) as follows:

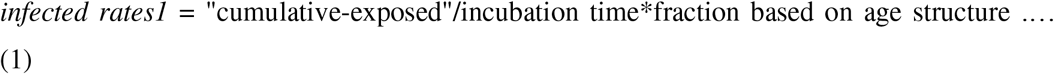

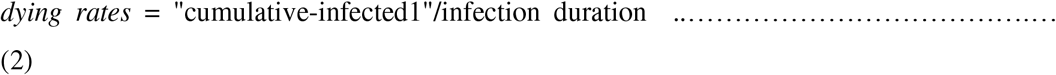

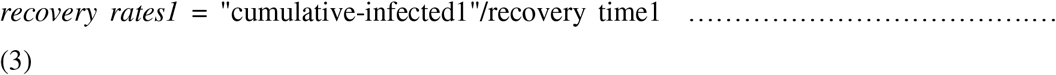

Transmission rate measures the number of exposed people after contacting or stand closes with infected people. Following Fiddaman (2020), transmission rate is calculated based on equation 3. For the first policy, its impact is calculated based on equation 4. Equation 4 means that the first policy of behavioral reduction risk decreases transmission rate based on two factors: “behavioral reduction risk” and “behavioral reduction time”.

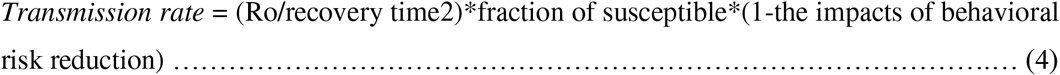

A similar equation is applied for the second policy as seen in equation 5. Equations 5a and 5b show the number of exposed cases decreases after the second policy starts at “lockdown risk reduction time”.

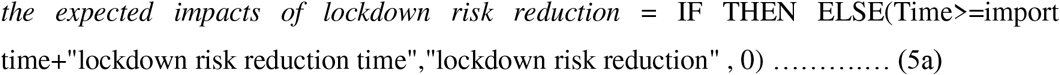

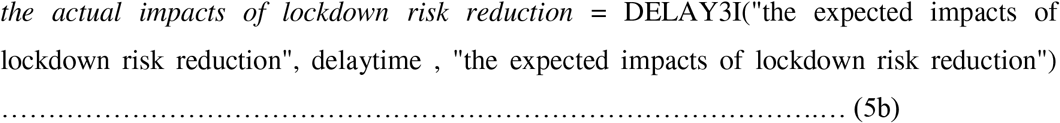

After running MCMC calibration process, the author finds some possible values with the smallest errors as seen in table 2.

**Table 2.** Best parameter values of the SEIR model. In bracketed value ranges based on existing studies as seen in table 1. No bracketed values mean that best values are assumed.

| Variables | Estimated values |
| --- | --- |
| Ro (basic reproduction number) | 3.45 (2.4-3.8) |
| Incubation time (days) | 3 (2-7) days |
| Infection duration (days) | 64.5 (45-91) days |
| Recovery time (days) |  |
| * recovery time1 ($\geq 60$ years) | 5 (3-35) days* |
| ** recovery time2 ($< 60$ years) | 3.5 (3-7) days** |
| Fraction based on age structure | 22% |
| The behavioral reaction time (days) | 15 |
| The behavioral risk reduction | 10% |
| The lockdown risk reduction time | 61 |
| The lockdown risk reduction | 84.5% |
| Delay time (days) | 4.5 |

It is found that the first policy efficacy is about 10%, assuming that an incubation time is about 3 days. While the second policy efficacy is about 85% in minimizing the COVID-19 flow. This finding may explain the second wave of the COVID-19 in Germany. Once the second policy i.e. the lockdown(s) are released the infected cases have risen significantly despite the public awareness or public behavior existence (wearing masks and social distancing in public spaces).

It appears that the SD model can reproduce similar outputs compared to respective observed outputs as seen in figure 2. The SD model performance shows that the SD model has symmetric Mean Percentage Errors (sMAPE) less than 10% (table 3).

**Table 3.** sMAPE for the SEIR model

| Variables | sMAPE |
| --- | --- |
| Infected cases | <5% |
| Death cases | <5% |
| Recovered cases | 7% |

## 4. Conclusions

This study fills a gap in which existing studies have not separated the impacts of the first policy (the people behavior) and the second policy (the government actions) in tackling the COVID-19 flow in Germany. To measure the impacts of the first and the second policy, this study develops the SEIR model based on the system dynamics approach and obtain the best parameter values through MCMC calibration process.

It is shown that the first policy i.e. preventive behavior such as handwashing and voluntarily social/physical distancing is important in minimizing the flow of the COVID-19. However, the second policy i.e. lockdowns shows is important roles in flattening the COVID-19 curve. With keep this point in mind, releasing lockdowns should be initiated carefully as, despite the importance of the preventive behavior, the first policy is not sufficient to significantly decrease the pandemic cases.

This study shows that the cohort SEIR model can successfully reproduce similar patterns of the COVID-19 in Germany. As it is the cohort SEIR model, it can be used to estimate the imbalance effects of the COVID-19 on different age groups. Moreover, as the cohort SEIR model can be a basis to simulate impacts of the COVID-19 on resources such as health workers and bed capacities.

## Data Availability

data is available in model dataset:germany_model_ref.vdf

## Supplemental material(s)

The SEIR model is available at: https://osf.io/3k6db

## Notes

### Competing Interest Statement

The authors have declared no competing interest.

### Funding Statement

There is no given funding

### Author Declarations

no ethical needed

### Summary of Updates

I did little revision of the abstract, figure 2, and table 3. I also did a little change of the title

